# Programmes evaluated after teaching infection prevention and control training in health or social care settings: A mapping review

**DOI:** 10.1101/2024.04.26.24306446

**Authors:** Julii Brainard, Isabel Catalina Swindells, Charlotte Christiane Hammer, Emilio Hornsey

## Abstract

**Objective:** To provide an overview of country settings, study designs, pathogens, response stage, outcomes and monitoring periods that were described in studies that may provide evidence about effectiveness of training in infection prevention and control programmes, for health or social care workers.

**Methods:** A systematic review was undertaken to find and summarise aspects of relevant studies published from 2000-2023. Eligible studies had to have pre and post evaluation or post-intervention evaluation in case of trials only. Eligible outcomes were knowledge; adherence/compliance; skills or practice; incidence; case-related mortality. Eligible infectious diseases were those caused by any single cell biological entity (eg virus or protozoa) where vectors were not the primary transmission pathway. Infection prevention settings had to be health/social care (not community or environmental), and participants had to be health or social care staff or trainee staff. Articles from three bibliographic databases were dual-screened independently and key data were extracted and verified. Findings are summarised quantitatively and narratively.

**Findings:** Included studies numbered 210, of which 187 were pre-post study design and 23 had concurrent comparator arms. Most studies (n=128) were undertaken in high income country settings, especially in the USA (n=31), and 47 were in European Union member countries. There were 20 studies based in China, and 5 in India. Frequency of phases were preparedness (n=47), readiness (n=29), response (n=146), and recovery (n=4). The most commonly mentioned pathogens were SARS-CoV-2 (n=73) and anti-microbial-resistant organisms (AMROs, n=54). Most settings were health care centres but long-term care facilities (n=13) and healthcare delivered by emergency responders (3) were also mentioned. Dental professionals or students were in just 3 studies and 10 studies had trainee health professionals as participants.

**Conclusion:** The research questions for which the most evidence is likely to exist about effectiveness of IPC training of health care workers would be in response phase in high income countries, especially if the relevant pathogens were AMROs or SARS-CoV-2. In contrast, the prospects are not good for finding evidence that could deliver confident conclusions about optimal IPC training programmes in low income countries, for most specific diseases (eg. cholera or tuberculosis) or in non-response phases.

## Introduction

Infection prevention and control (IPC) is a routine and essential activity in health and social care environments. In spite of many initiatives in recent years to develop national IPC training guidelines (3, 4), before the Covid-19 pandemic many health and social care workers (HCWs) globally reported that they had received little training in IPC (5). It now seems likely that most HCWs who worked in the 2020-2023 period received at least some formal IPC training, but how often that training was effective or met objective standards is unclear. The effectiveness of the specific teaching formats and methods to train HCWs in IPC have not previously been evaluated thoroughly and systematically, to identify which curricula or pedagogical elements might be consistently successful. A systematic review to identify which teaching formats and methods were most applied and potentially replicable in fragile, vulnerable or conflict-affected (FCV) settings during response or readiness phases (Brainard et al. 2024 under review) concluded that most such IPC training programmes were not described adequately to be replicable and were additionally likely to suffer from many biases that undermined confidence in their apparent success, such as unclear attrition, selective recruitment and reporting and lack of standardised instruments to capture outcomes. Controlled trial study designs were unusual and lack of unsuccessful interventions in the available reports suggested publication bias (6).

We undertook and report here a mapping review (7) which aimed to systematically collect and report information that describes the potential evidence base for evaluation of IPC training methods and formats for HCWs, with respect to key contextual factors. A mapping review may be distinguished from scoping or systematic reviews in that a mapping review typically does not synthesise evidence in favour of or against any specific intervention, but rather reports information preliminary to understanding which research questions are viable to explore. For instance, with regard to IPC training, a mapping review could suggest if many studies exist that describe IPC training to prevent a specific disease (e.g., cholera or norovirus), in a specific context (eg. low income countries), provide information about specific outcomes (eg. knowledge change or handwashing compliance) or if training was delivered during a specific response phase (such as preparedness, when an known active infectious threat is not concurrently present (8)). Most health-related systematic reviews on effectiveness are focused by constraining inclusion and exclusion criteria related to intervention format, disease process, situational context and/or specific possible benefits or harms (9).

## Methods

Our PROSPERO registration number is CRD42023472400. Deviations from protocol were that we did not use the Covidence platform and we did not undertake backwards or forwards citation searches or quality assessment or extract information on secondary outcomes. This article is one of several outputs arising from the same protocol.

### Searches

MEDLINE, Scopus, LILACS were searched on 9 October 2023 with the phrase (Scopus syntax):

(“infection-control”[Title/Abstract] or “transmission”[Title/Abstract] or “prevent-infectio*”[Title/Abstract])

And

(“emergency”[Title/Abstract] or “epidemic”[Title/Abstract] or “outbreak”[Title/Abstract]) and

(“training”[Title/Abstract] or “educat*”[Title/Abstract] or “teach*”[Title/Abstract])

Included studies in three recent and highly relevant systematic reviews (1, 2) were also screened. We only searched for studies in peer-review (scientific) literature.

### Screening and inclusion criteria

All studies found by the search strategy were independently screened by two authors, with decisions recorded on MS Excel spreadsheets. A single author (JB) revisited all initial choices to make final decisions. The final decision about inclusion was made from the abstract if the abstract contained sufficient information, else the full text was consulted. There were no language exclusions, but studies without a coherent abstract in English (available from the bibliographic database or using Google translate) were treated as unavailable for assessment.

Participants had to be health care or social care professionals or trainees in health or social care working in a clinical or residential care setting. We referred to World Health Organisation occupational definitions (10) if in doubt. Volunteer participants such as peer leaders or family carers were excluded. Study designs were either trials or pre-post. Pre-post studies had to report both baseline and post-training measurements of a primary outcome; only post-training outcomes had to be reported in trials.

To focus on relatively modern methods and understanding, studies were excluded if published before 2000. Infections from complex multicellular organisms (like human mites or lice) were excluded. Our focus was on IPC training related to ongoing individual action that should result in relatively quick benefits, in clinical, not community, environments. We therefore excluded interventions or outcomes that related to screening, surveillance, vaccination programmes, forms of patient care (e.g., Anti-viral treatment) that might reduce infectious period, availability of PPE or other resources that reflect institutional will and opportunity as much as any individual action, testing or screening strategies, or protocols or actions to hasten test results. All of these can reduce infection prevalence, but are outside the traditional understanding of IPC training. Interventions that primarily addressed environmental management outside of the clinical/care environment were also excluded, with exception of programmes that addressed clinical waste generated and managed on site.

Although we did not assess effectiveness in this study, our objective was to inventory the evidence base that might provide information about specific primary outcomes and therefore we imposed the requirement that included studies had to report at least one of the primary outcomes. We excluded studies when the primary outcomes were only measured > 12 months after training started (eg., quality improvement projects) because of the complexities that arise in trying to confidently associate exposure and outcomes over a long period. We also exclude cross-sectional studies without baseline assessment of pre-training outcomes.

### Data extraction, coding and summary

Fields extracted for each study were: bibliographic details, country (or countries) where the training was delivered; phase (8) that training was delivered in (preparation, readiness, response, recovery); study design; clinical environment as described; primary outcomes reported; time from intervention end to outcome assessment. Countries were categorised according to four World Bank income groups (2022 classification; 11) as high income, upper or lower middle income, or lower income. We summarise the results broadly and narratively.

## Results

Figure 1 shows the study selection process. 210 studies were included. Extracted data and bibliographic details are in a supplemental file available from authors (S1). Most studies (n=187) were pre-post design; 23 were experimental studies (controlled trials). Table 1 lists summary information about the included studies. A small number of studies didn’t fit into the categories in Table 1. For instance, two studies (12, 13) described training delivered by militaries of high income countries for IPC implementation in multiple other countries (not high income). Most clinical settings were described only generically, as hospitals or clinics or “health care facilities”. Most were relatively large medical centres described as hospitals or single hospital departments. The most mentioned specialisms described specifically were intensive care units (n=26) and neonatal or paediatric care units (n-19). Other specialisms which were each mentioned one to four times were: dental students or professionals, cardiac units, surgical units, burns treatment, urology centre, cystic fibrosis centre, emergency departments and emergency responders. Ten studies reported training that was delivered to trainees or students.

**Table 1.** Counts (%) of included studies in each category.

| *Attribute* |  |  |  |  |
| --- | --- | --- | --- | --- |
| **World Bank country income group** | High<br>128 (61%) | Upper middle<br>41 (20%) | Lower middle<br>32 (15%) | Lower<br>7 (3%) |
| **Response stage** | Prep.<br>47 (22%) | Readiness<br>29 (14%) | Response<br>146 (70%) | Recovery<br>4 (2%) |
| **Disease** | AMRO<br>54 (26%) | Covid-19<br>73 (33%) | Not specific<br>40 (19%) | EbD<br>16 (8%) |
| **Context or setting** | Trainees<br>10 (5%) | Clinical<br>187 (89%) | Long term care facility<br>13 (6%) |  |
| **Counts of studies with specific outcomes** | Case#/M<br>113 (54%) | Knowledge<br>60 (29%) | Skills or practice<br>30 (14%) | Compliance, behaviour or adherence<br>49 (23%) |
| **Counts of studies where the outcomes were assessed only immediately** | Case#<br>0 | Knowledge<br>27 (45% of 60) | Skills or practice<br>14 (47% of 30) | Adherence, behaviour or compliance<br>6 (12% of 49) |

**Notes:** Prep. = preparedness stage; in some cases, the training was delivered in multiple phases/settings/countries or addressed multiple specific diseases. EbD = Ebola virus disease. Case#/M = Case counts/incidence or related-mortality counts/incidence. AMRO = anti-microbial-resistant organism.

**Figure 1.**
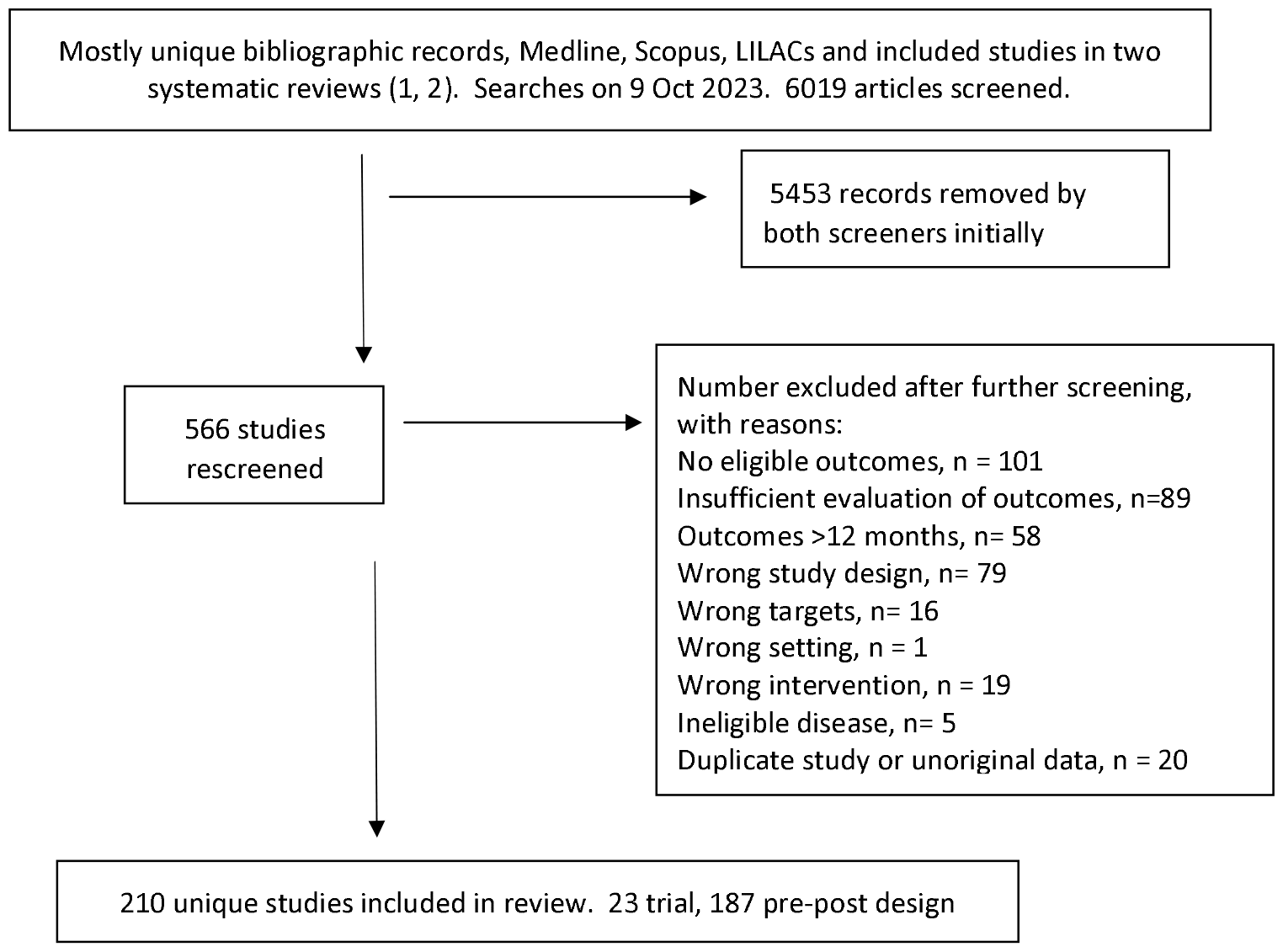
Selection procedure *Note*: More than one reason for exclusion was sometimes recorded.; not every reason for exclusion was recorded.

Most training was delivered in the response phase (n=146, 70%), when there was an active disease threat. About 39% (n=57/146) of the IPC training delivered in the response phase related to Covid-19. Anti-microbial resistant organisms were specific targets in about 25% (n=37) of the 146 studies delivered during response phase. A large variety of other specific diseases were mentioned such as pertussis, norovirus, tuberculosis, varicella, cholera, etc., each in fewer than 5 studies.

Case counts (n=115) were the most commonly reported patient outcomes; mortality was reported in just 4 studies. Knowledge (n=60) was the most reported of the outcomes in health care workers that most directly relate to personal capacity. A majority of studies (between 55% and 100%, depending on outcome) had assessment points beyond the immediate finish after training.

## Discussion

This review finds that the most viable research questions to explore with regard to effectiveness of IPC training would be in response phases in clinical settings in high income countries, with outcomes to consider being case counts, knowledge or possibly adherence domains, and especially if faced with AMROs or Covid-19. Most studies had assessment points after the training finished. For instance, we found 43 studies that assessed compliance or adherence behaviours more than once after training completed; a later assessment point is optimal to support conclusions about whether taught IPC practices became habitual. In contrast, the prospects are not good of finding evidence that could deliver confident conclusions about best design for IPC training programmes in low income countries, for most specific diseases (eg., cholera or tuberculosis), or in the recovery phase (only 4 studies described training in the recovery context). Our results suggest that a systematic evidence synthesis evaluating effectiveness of IPC training with inclusion criteria imposed about most specific possible settings, any single outcome and most possible specific infectious diseases, would be unlikely to find more than 5-10 primary research studies.

That count (5-10) has bearing on the feasibility of obtaining conclusive results in an evidence overview. How many studies are required to undertake evidence synthesis that yields highly certain results is a moot point. This topic is large and arguably rapidly developing, and thus outside our scope to thoroughly explore. We provide only some suggestive guidance here. How many studies are enough in an evidence synthesis at least partly depends on which sources of bias or uncertainty are deemed to be most important, and which methods are used to synthesise or describe the consistency and certainty of the evidence. Study design itself is important, in that randomised controlled trials are widely considered to be a least biased primary interventional study design (14); we note that relatively few trials were found in our evidence search. Quantitative primary study outcomes are often pooled using meta-analysis. A commonly quoted metric for describing heterogeneity between studies in meta-analysis results is the I^2^ statistic; it has been suggested that if I^2^ is applied to < 15 studies, the proportion of variability in a meta-analysis may be better explained not by sampling error but rather by other differences between the included trials (15); these other differences undermine confidence in obtaining similar results in a new study. With regard to capturing trends between studies, meta-regression is an umbrella term for a set of evidence synthesis methods (16) that describe correlation between outcomes and study-level variables. Meta-regression can thus be used for trend analysis or to look for consistency of association. The Cochrane handbook suggests that meta-regression should not be applied for fewer than ten studies (17). The GRADE (Grading of Recommendations Assessment, Development and Evaluation) (18) approach is a commonly applied and well-developed framework for estimating certainty in effect size arising from meta-analysis and other evidence synthesis exercises (19). GRADE does not explicitly incorporate count of included studies as part of estimating certainty, but it does look for bias in many domains in the primary studies and the grouped evidence. This means that, in practice, outcomes need to be reported from a relatively large number of studies for a high GRADE assessment to be assigned.

Strengths of our review include searches in multiple languages and three bibliographic databases. The search strategy had high sensitivity (we rescreened all studies with at least one initial screener vote) and a large sample size was achieved. Our review also had defined limitations. We confined eligible outcomes to knowledge, skills or practice, adherence/compliance, and case incidence or mortality. These outcomes were chosen on the basis of preliminary literature searches about frequency of evaluated outcomes and our own judgement about outcomes that could plausibly relate to IPC training. They aren’t the only outcomes of interest nor necessarily the best outcomes to consider. Evaluation of incidence and mortality are especially problematic indicators of IPC training success, because they are not necessarily direct benefits from good IPC practices. Incidence often depends highly on local community prevalence of relevant pathogen(s), while mortality rates often reflect quality of health care available in addition to timing of care seeking.

This review cannot comment about many other key factors that can be important to intervention effectiveness. Two important intervention aspects that we did not address are teaching methods and formats (20), and whether comprehensive guidelines were used to design the appropriate training curriculum (3). We also did not look for process evaluation information (21). Process evaluation can be essential to understanding why any particular behavioural intervention was successful, but most interventions were probably not subject to process evaluation.

The Covid-19 pandemic probably accelerated development of standardised IPC training programmes in many settings and for many national workforces (5). However, in 2020-2023 many such training programmes were developed quickly and delivered without evaluation of effectiveness or with only pre-post study design evaluation. We have shown that there is substantial opportunity to design appropriate randomised trials to confidently identify the most effective forms of IPC training for health and social care staff. In an effort to understand the effectiveness of different dissemination strategies, Silva *et al*. 2021 (2) undertook a systematic review to find randomised controlled trials intended to improve adherence with IPC guidelines on preventing respiratory infections in healthcare workplaces. The Silva et al. review included 14 interventions, all but one in high income countries (in Iran; 22). Their result suggests that the greatest opportunities for knowledge gains in improving IPC training delivery may arise from delivering trials in lower and middle income countries.

## Data Availability

All data produced in the present study are available upon reasonable request to the authors

## Author contributions

Analysis plan: JB, CCH

Comments on draft manuscript: JB, ICS

Conception: JB, CCH

Data acquisition and extraction: JB, ICS

Data curation: JB

Data synthesis: JB

Funding: JB, CCH

Interpretation: All authors

Research governance: JB

Screening: JB, ICS, CCH

Writing first draft, assembling revisions: JB

## Acknowledgements

Comments and advice from Joanna Wild (OL4All, Oxford UK), and World Health Organization (WHO) staff Victoria Willet and Hibak Mahamed and an expert working group coordinated by the WHO all shaped the study protocol and methods.

## References

1. Nayahangan LJ, Konge L, Russell L, Andersen S. Training and education of healthcare workers during viral epidemics: a systematic review. BMJ Open. 2021;11(5):e044111.

2. Silva MT, Galvao TF, Chapman E, da Silva EN, Barreto JOM. Dissemination interventions to improve healthcare workers’ adherence with infection prevention and control guidelines: a systematic review and meta-analysis. Implementation Science. 2021;16(1):1–15.

3. Qureshi MO, Chughtai AA, Seale H. Recommendations related to occupational infection prevention and control training to protect healthcare workers from infectious diseases: a scoping review of infection prevention and control guidelines. BMC Health Services Research. 2022;22(1):272.

4. Barratt R, Gilbert GL. Education and training in infection prevention and control: exploring support for national standards. Infection, Disease & Health. 2021;26(2):139–44.

5. Tartari E, Tomczyk S, Pires D, Zayed B, Rehse AC, Kariyo P, et al. Implementation of the infection prevention and control core components at the national level: a global situational analysis. Journal of Hospital Infection. 2021;108:94–103.

6. Song F, Hooper L, Loke YK. Publication bias: what is it? How do we measure it? How do we avoid it? Open Access Journal of Clinical Trials. 2013;5:71.

7. Campbell F, Tricco AC, Munn Z, Pollock D, Saran A, Sutton A, et al. Mapping reviews, scoping reviews, and evidence and gap maps (EGMs): the same but different—the “Big Picture” review family. Systematic Reviews. 2023;12(1):45.

8. WHO. Emergency cycle. World Health Organization; 2024 [cited 2024 Jan 9]; Available from: https://www.who.int/europe/emergencies/emergency-cycle.

9. Stern C, Jordan Z, McArthur A. Developing the review question and inclusion criteria. American Journal of Nursing. 2014;114(4):53–6.

10. WHO. Health Workforce-related terminology: Terminology work carried out by the WHO Language department at the request of the Health Workforce department. 2021 [updated Jun 8; cited 2024 Jan 9]; Available from: https://cdn.who.int/media/docs/default-source/health-workforce/hwp/202100608-health-workforce-terminology.pdf.

11. World Bank Blogs. New World Bank country classifications by income level: 2022-2023. World Bank; 2023 [cited 2024 Jan 10]; Available from: https://blogs.worldbank.org/opendata/new-world-bank-country-classifications-income-level-2022-2023.

12. Hsu C-C, Tsai S-H, Tsai P-J, Chang Y-C, Tsai Y-D, Chen Y-C, et al. An Adapted Hybrid Model for Hands-On Practice on Disaster and Military Medicine Education in Undergraduate Medical Students During the COVID-19 Pandemic. Journal of Acute Medicine. 2022;12(4):145.

13. Crouch HK, Murray CK, Hospenthal DR. Development of a deployment infection control course. Military Medicine. 2010;175(12):983–9.

14. Petrisor B, Bhandari M. The hierarchy of evidence: levels and grades of recommendation. Indian Journal of Orthopaedics. 2007;41(1):11.

15. Thorlund K, Imberger G, Johnston BC, Walsh M, Awad T, Thabane L, et al. Evolution of heterogeneity (I2) estimates and their 95% confidence intervals in large meta-analyses. PloS One. 2012;7(7):e39471.

16. Thompson SG, Higgins JP. How should meta-regression analyses be undertaken and interpreted? Statistics in Medicine. 2002;21(11):1559–73.

17. Cumpston M, Li T, Page MJ, Chandler J, Welch VA, Higgins JP, et al. Updated guidance for trusted systematic reviews: a new edition of the Cochrane Handbook for Systematic Reviews of Interventions. The Cochrane Database of Systematic Reviews. 2019;2019(10).

18. Hultcrantz M, Rind D, Akl EA, Treweek S, Mustafa RA, Iorio A, et al. The GRADE Working Group clarifies the construct of certainty of evidence. Journal of Clinical Epidemiology. 2017;87:4–13.

19. Murad MH, Mustafa RA, Schünemann HJ, Sultan S, Santesso N. Rating the certainty in evidence in the absence of a single estimate of effect. BMJ Evidence-Based Medicine. 2017.

20. Barrera-Cancedda AE, Riman KA, Shinnick JE, Buttenheim AM. Implementation strategies for infection prevention and control promotion for nurses in Sub-Saharan Africa: a systematic review. Implementation Science. 2019;14(1):1–41.

21. French C, Pinnock H, Forbes G, Skene I, Taylor SJ. Process evaluation within pragmatic randomised controlled trials: what is it, why is it done, and can we find it?—a systematic review. Trials. 2020;21(1):1–16.

22. Jeihooni AK, Kashfi SH, Bahmandost M, Harsini PA. Promoting preventive behaviors of nosocomial infections in nurses: The effect of an educational program based on health belief model. Investigacion y Educacion en Enfermeria. 2018;36(1).

